# THE GENETIC LANDSCAPE OF HEREDITARY SPASTIC PARAPLEGIA IN CANADA

**DOI:** 10.64898/2026.10.14.26365432

**Authors:** Sylvine Niyoyita, Mehrdad A. Estiar, Han Yu, Eric Yu, Saeid Amiri, Jamil Ahmad, Sian D. Spacey, Kym M. Boycott, Mark Tarnopolsky, Oksana Suchowersky, Grace Yoon, Nicolas Dupré, Guy A. Rouleau, Ziv Gan-Or

**Affiliations:** Department of Human Genetics, McGill University, Montréal, QC, Canada; The Neuro (Montreal Neurological Institute-Hospital), McGill University, Montréal, QC, Canada; Department of Quantitative Life Science, McGill University, Montréal, QC, Canada; Department of Neurology and Neurosurgery, McGill University, Montréal, QC, Canada; Division of Neurology, Department of Medicine, University of British Columbia, Vancouver, British Columbia, Canada; UBC Hospital and Vancouver General Hospital, Vancouver, British Columbia, Canada; CHEO Research Institute; University of Ottawa, Ottawa, ON, Canada; Department of Pediatrics, McMaster University, Hamilton, ON, Canada; Department of Medical Genetics, Faculty of Medicine & Dentistry, University of Alberta, Edmonton, AB, Canada; Divisions of Neurology and Clinical and Metabolic Genetics, Department of Paediatrics, The Hospital for Sick Children, University of Toronto, Toronto, ON, Canada; Department of Medicine, Faculty of Medicine, Université Laval; Neuroscience Axis, CHU de Québec–Université Laval Research Centre, Québec City, QC, Canada; Broad Institute of Harvard and MIT, Cambridge, MA, USA

## Abstract

**Background:** Hereditary spastic paraplegia (HSP) comprises a heterogeneous group of neurodegenerative disorders characterized by progressive lower-extremity weakness and spasticity, with or without additional neurological symptoms.

**Objectives:** To characterize the molecular genetic spectrum, inheritance patterns, molecular finding rate, and clinical characteristics of HSP within a multicenter Canadian cohort.

**Methods:** We investigated 671 patients from 423 families who met clinical criteria for HSP identified through the Canadian HSP (CanHSP) registry from 7 medical centers across Canada. Patients were evaluated using known HSP and HSP mimic gene panels, whereas unsolved and more recent patients underwent whole-exome sequencing (WES). 435 individuals underwent gene-panel testing and 485 underwent WES. Copy-number variants (CNVs) were assessed using ExomeDepth, and candidate CNVs using multiplex ligation-dependent probe amplification (MLPA).

**Results:** Molecular findings were identified in 426 of 671 individuals (63.5%): 351 (52.3%) had HSP-associated findings, 70 (10.4%) HSP-mimicking disorders, and five (0.7%) other molecular findings; 245 (36.5%) remained unresolved. Candidate variants were found in 80 relevant genes, 36 of which were HSP-associated genes. The most frequently implicated genes were *SPAST* (22.5%), *SPG7* (5.7%), *ATL1* (5.2%), *SPG11* (3.6%), and *SPG8* (3.3%). Phenotypically, *SPAST*, *ATL1*, and *SPG8* were predominantly associated with pure HSP, whereas *SPG7* and *SPG11* more often showed complicated phenotypes, with prominent cerebellar involvement in *SPG7* and cognitive and developmental features in *SPG11*.

**Conclusion:** Patients in CanHSP present with genetic and phenotypic heterogeneity. Reanalysis and complementary approaches, including whole-genome, RNA, and long-read sequencing, may increase diagnostic yield.

## 1. Introduction

Hereditary spastic paraplegia (HSP) constitutes a group of heterogeneous neurodegenerative disorders characterized by progressive lower-extremity weakness and spasticity, with or without additional neurological symptoms ^1–3^. Although individually rare, HSPs are an important cause of lifelong motor disability ^4,5^. Clinically, HSP is classified as pure or complicated. In the pure form, symptoms are confined to lower-extremity spasticity and weakness, whereas complicated HSP includes additional neurological and systemic manifestations such as ataxia, cognitive impairment, epilepsy, peripheral neuropathy, and extrapyramidal disorders ^4,6^.

Over the past two decades, substantial progress has been made in elucidating the genetic basis of HSP, with more than 90 disease-associated genes nominated ^4,6,7^. Many of the earliest discoveries, including genes responsible for autosomal dominant and autosomal recessive forms, predated the clinical adoption of next-generation sequencing ^8^. Since the introduction of whole-exome sequencing, its use has expanded the spectrum of detectable variants and facilitated molecular diagnosis in diverse populations ^1,9–12^.

Nevertheless, some challenges persist. A considerable proportion (30%-60%) of affected individuals remain genetically undiagnosed after comprehensive exome-based testing ^10,12–14^. This diagnostic gap likely reflects multiple factors, including undetected structural variants, noncoding or regulatory mutations, limitations in variant interpretation, and the contribution of yet-unidentified disease genes ^13,15^. Furthermore, the relative contribution of HSP-associated genes appears to vary across populations ^5,16–18^, emphasizing the importance of large, well-characterized national cohorts.

The Canadian population offers a unique setting to examine the genetic landscape of HSP due to its ethnic diversity, regional founder effects, and centralized academic referral networks ^5,19^. In this study, we investigated a Canadian HSP (CanHSP) cohort reflecting the evolving diagnostic practices, adding to a series of studies characterizing HSP genetics in Canada from the CanHSP group ^5,14,16,20,21^. We sought to characterize the molecular genetic spectrum of HSP, quantify the proportion of genetically resolved and unresolved cases, define the relative contribution of the most frequently implicated genes in this population, and examine the phenotypes associated with common genetic causes of HSP.

## 2. Methods

### 2.1. Population

We investigated 671 patients with suspected HSP from 423 families enrolled in CanHSP, a multicenter registry maintained by seven medical centers in Quebec, Alberta, British Columbia, and Ontario. The geographic distribution of the HSP cohort across Canada is shown in Supplementary Figure S1. The earliest patient evaluation was in 1979. In total, 435 individuals underwent gene-panel testing and 485 underwent WES; because some individuals underwent both approaches, these categories were not mutually exclusive. The overall testing pathway and molecular outcome are summarized in Supplementary Figure S2. Clinical data were updated annually when available, with the last available update ranging from 2012 to 2022. All participants provided written informed consent, and all data were stored in a central database at McGill University. The study was approved by the McGill University Research Ethics Board. Demographic and clinical variables collected included sex, age at onset, age at evaluation, family history, molecular testing results, developmental and cognitive features, core neurologic signs, associated neurologic symptoms, and severity scores (the Spastic Paraplegia Rating Scale [SPRS] and SPATAX-EUROSPA disability score), and walking aid use.

Phenotype was classified as pure or complicated HSP. Pure HSP was defined as predominantly isolated lower limb spasticity with or without weakness and urinary symptoms, whereas complicated HSP was defined by the presence of additional neurologic or systemic features beyond the core syndrome, including progressive cognitive deficits, retinopathy/optic atrophy, seizures, upper- or lower-extremity ataxia, peripheral neuropathy, dysarthria, deafness, skeletal abnormalities, or myoclonus.

### 2.2. Molecular Genetic Analysis

To determine the molecular basis of disease in CanHSP, we performed genomic analysis using standard DNA extraction ^22^ and sequencing procedures. Initially, gene-panel testing was used as the diagnostic approach. Patients who remained genetically unsolved after panel testing, as well as more recent cases, underwent whole-exome sequencing (WES). For patients undergoing WES, exome capture and target enrichment were performed using the Agilent SureSelect Human All Exon v4 kit. Samples were sequenced at the Genome Québec Innovation Centre (Montreal, QC, Canada) on the Illumina HiSeq 2000/2500 platform. We aligned sequencing reads to the human reference genome (GRCh37) using Burrows-Wheeler Aligner (BWA) ^23^, performed variant calling using the Genome Analysis Toolkit (GATK) ^24^, and performed functional annotation using ANNOVAR ^25^. Exome data were screened for genes known to cause HSP as well as genes associated with overlapping neurogenetic disorders that may present with spastic paraplegia ^26,27^. Copy-number variants (CNVs) were assessed from exome sequencing data using ExomeDepth ^28^, and candidate CNVs were historically evaluated using the SALSA MLPA kit (MRC-Holland; Probemix P213 HSP mix-2)^29^. Historical CNV/MLPA findings were retained as molecular findings only when sufficient supporting documentation was available to confirm the molecular alteration, including the relevant exon-level or genomic information. Interpretation was established in concordance with clinical presentation and literature and classified as HSP or HSP mimics. Genes associated with hereditary spastic paraplegia, including genes with established SPG designations or well-described HSP phenotypes, were classified as HSP-associated genes. Genes primarily associated with other neurogenetic disorders were classified as HSP mimics when the associated disorder was known to include progressive lower-extremity spasticity, spastic paraparesis, spastic gait, corticospinal tract signs, or an overlapping phenotype, and when the patient’s recorded clinical features were compatible with that disorder ^30,31^. For disorders with partial clinical overlaps, classification required genotype-phenotype concordance; genes whose associated conditions did not plausibly explain the recorded HSP-like phenotype were not considered definitive mimic findings. For patients with more than one molecular finding, each individual was assigned once to a final molecular outcome category (HSP-associated, HSP-mimic, or other) according to the finding most consistent with the recorded phenotype, inheritance pattern, and known disease mechanism. For gene-level frequency analyses, however, every implicated gene was counted; therefore, gene-level counts were non-mutually exclusive. The list of analyzed genes is provided in Supplementary Table S4. Mode of inheritance was assigned according to available pedigrees and the identified genes. Patients and families were categorized as autosomal dominant (AD), autosomal recessive (AR), X-linked, and unknown, when applicable.

#### Variant Assessment and Interpretation

Variants were assessed from registry records, according to the American College of Medical Genetics and Genomics and the Association for Molecular Pathology (ACMG/AMP) criteria ^32^ through Franklin/Genoox ^33^, variant databases and published literature where available.

To assess variant novelty, candidate variants were annotated using ANNOVAR against population and variant databases, including the Genome Aggregation Database (gnomAD) v2.1.1, the Exome Aggregation Consortium (ExAC), the 1000 Genomes, the Single Nucleotide Polymorphism Database (dbSNP), and ClinVar. Variants absent from these resources were reviewed in HGMD Professional and the Leiden Open Variation Database (LOVD), and by manual literature searches using gene symbols, HGVS cDNA and protein nomenclature, and hg19 genomic coordinates. Variants absent from population databases, disease databases, and the published literature were considered candidate novel variants. Novelty was interpreted separately from pathogenicity.

### 2.3. Statistical analysis

To describe the cohort and compare major genetic and clinical subgroups, we performed descriptive and comparative statistical analyses in the full dataset. Continuous variables were summarized using median and interquartile range (IQR), with mean and standard deviation provided as supplementary descriptive summaries where relevant. Categorical variables were summarized using frequencies and percentages. Clinical variables were analyzed using available data denominators.

Group comparisons for categorical variables were performed using Fisher’s exact test when expected cell counts were small; otherwise, chi-square tests were used. Mann–Whitney U test was applied for non-normally distributed continuous data. Variables examined included sex, age at onset, age at evaluation, familial status, inheritance pattern, solved status, and available severity measures. Analyses were performed in R software (R v4.5.2) ^34^, and the analysis code was made publicly available on GitHub at: https://github.com/sylvineniy/HSP.

## 3. Results

### 3.1. Cohort composition and clinical characteristics

The cohort included 671 patients, with a slight male predominance (366/671, 54.5%), while 297/671 (44.3%) were female and 8/671 (1.2%) unspecified. Median age at onset was 15.5 years (IQR 2.0–37.0), and median age at last recorded evaluation was 40.0 years (IQR 21.0–57.0) (Table 1).

**Table 1.**
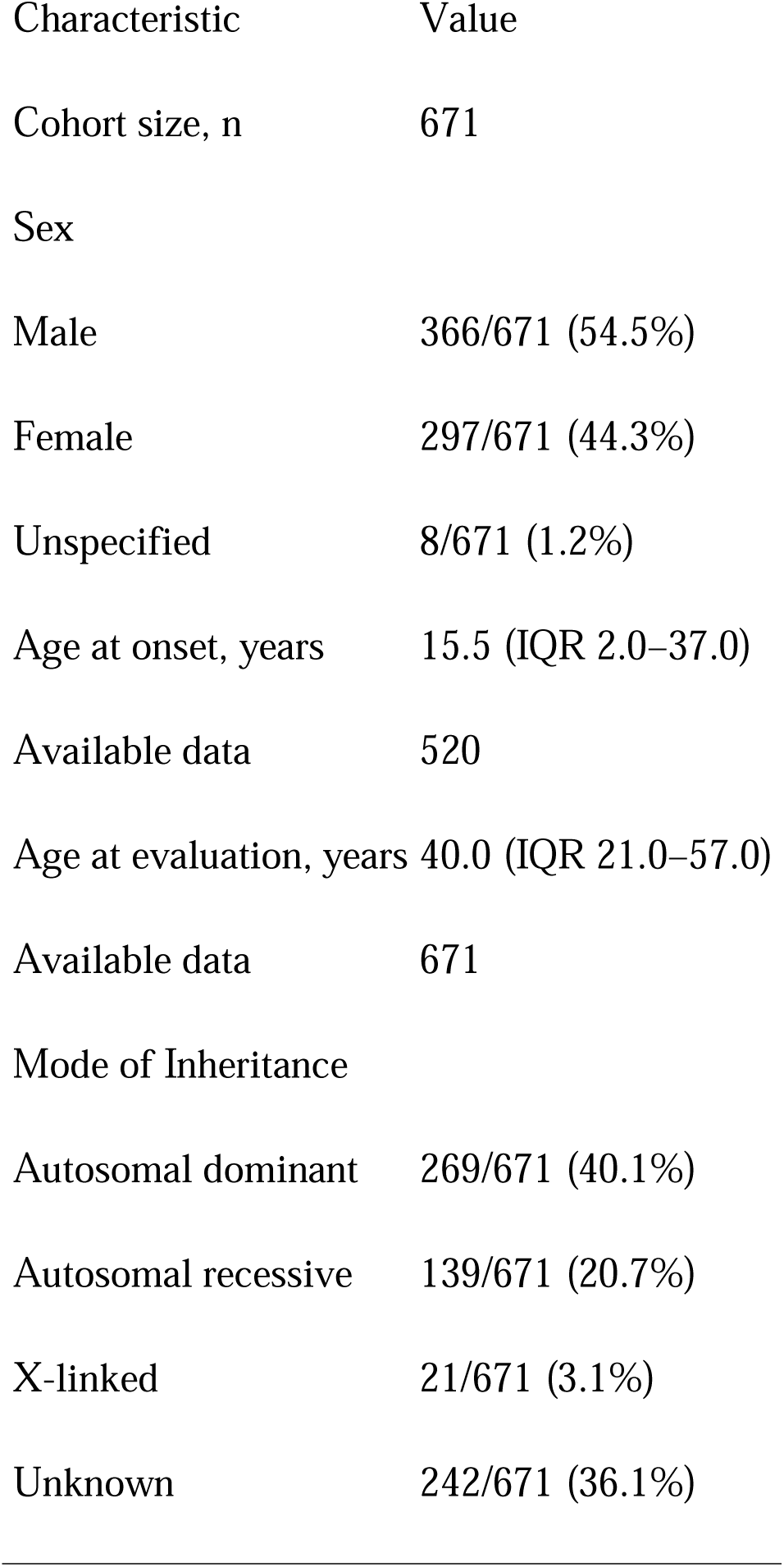
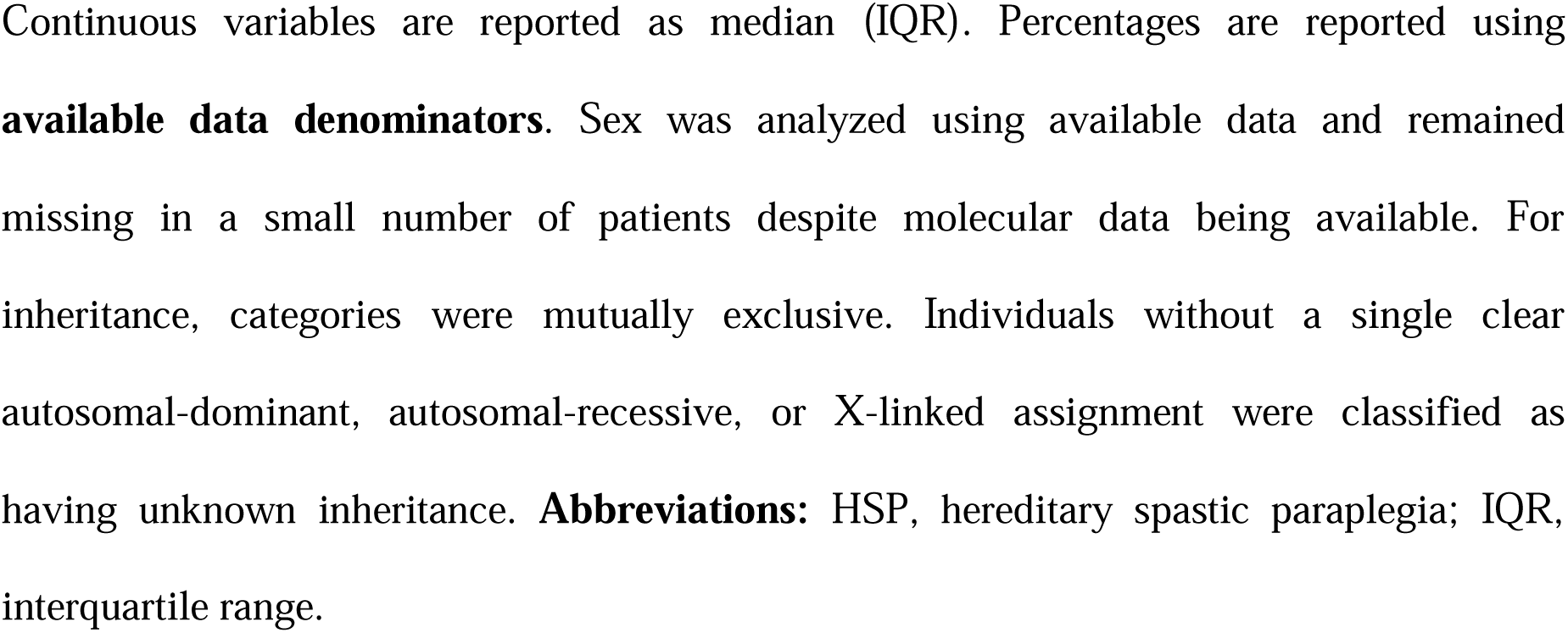
Clinical and demographic characteristics of the clinically ascertained HSP cohort.

| Characteristic | Value |
| --- | --- |
| Cohort size, n | 671 |
| Sex |  |
| Male | 366/671 (54.5%) |
| Female | 297/671 (44.3%) |
| Unspecified | 8/671 (1.2%) |
| Age at onset, years | 15.5 (IQR 2.0–37.0) |
| Available data | 520 |
| Age at evaluation, years | 40.0 (IQR 21.0–57.0) |
| Available data | 671 |
| Mode of Inheritance |  |
| Autosomal dominant | 269/671 (40.1%) |
| Autosomal recessive | 139/671 (20.7%) |
| X-linked | 21/671 (3.1%) |
| Unknown | 242/671 (36.1%) |

Continuous variables are reported as median (IQR). Percentages are reported using **available data denominators**. Sex was analyzed using available data and remained missing in a small number of patients despite molecular data being available. For inheritance, categories were mutually exclusive. Individuals without a single clear autosomal-dominant, autosomal-recessive, or X-linked assignment were classified as having unknown inheritance. **Abbreviations:** HSP, hereditary spastic paraplegia; IQR, interquartile range.

In patients with available phenotypic data, the most frequently recorded features were, as expected, lower-extremity spasticity, present in 444/520 patients (85.4%), lower-extremity hyperreflexia in 440/520 (84.6%), extensor plantar responses in 387/520 (74.4%), and lower-extremity weakness in 317/520 (61.0%, Supplementary Table S1). Other features included bladder dysfunction (197/520, 37.9%), upper-extremity hyperreflexia (173/492, 35.2%), sensory abnormalities (157/492, 31.9%), and dysarthria (133/492, 27.0%). Cerebellar and developmental features were less frequent, and seizures, retinopathy/optic atrophy, deafness, and myoclonus each affected fewer than 5% of patients with available data (Supplementary Table S1). Severity assessments, including SPRS, SPATAX-EUROSPA disability score, and walking-aid use, were available in subsets of patients and are summarized in Supplementary Tables S2 and S3.

Pure HSP had more frequent familial presentation (i.e., more than one patient with HSP in a pedigree) compared with complicated HSP (182/212 [85.8%] vs 62/103 [60.2%]; p<0.001). Inheritance pattern also differed by HSP type: pure HSP was predominantly autosomal dominant (179/239 [74.9%]), whereas complicated HSP showed nearly equal proportions of autosomal dominant (55/116 [47.4%]) and autosomal recessive inheritance (53/116 [45.7%]). X-linked inheritance was more frequent in complicated than pure HSP (8/116 [6.9%] vs 6/239 [2.5%]; p = 0.076). Complicated HSP showed greater disease severity and disability, with higher median SPRS scores (21 [15–31.5] vs 15 [8–22]) and higher median SPATAX-EUROSPA disability stages (3 [2–5] vs 2 [0–3]; both p<0.001).

### 3.2. Molecular findings and diagnostic architecture

Molecular findings were identified in 426 of 671 individuals (63.5%). These included 351 individuals (52.3%), representing 192 out of 423 families (45.4%), with HSP-associated molecular findings, and 70 individuals (10.4%) with molecular findings in genes associated with disorders that may clinically mimic HSP. Patient-level frequencies and clinical features for the 351 individuals with HSP-associated molecular findings are provided in Supplementary Table S5. Five individuals (5/671, 0.7%), representing 4 of 423 families (0.9%), had molecular findings in genes considered clearly outside established HSP-associated or HSP-mimicking disease categories in the available dataset (Supplementary Table S9). In addition, at the variant level, 71 unique sequence variants were classified as pathogenic or likely pathogenic, including 53 pathogenic and 18 likely pathogenic variants. Additional variants were classified as benign or variants of uncertain significance (VUS) and are summarized in Supplementary Table S6.

The remaining 245 patients (36.5%) had no molecular finding identified and were classified as unresolved. Nine patients had historical exon-level CNV findings recorded in the cohort; however, these findings could not be sufficiently confirmed from the available case-level documentation and were therefore not retained as molecular findings. The corresponding patients were classified as unresolved. The diagnostic architecture of the cohort is summarized in Figure 1.

**Figure 1.**
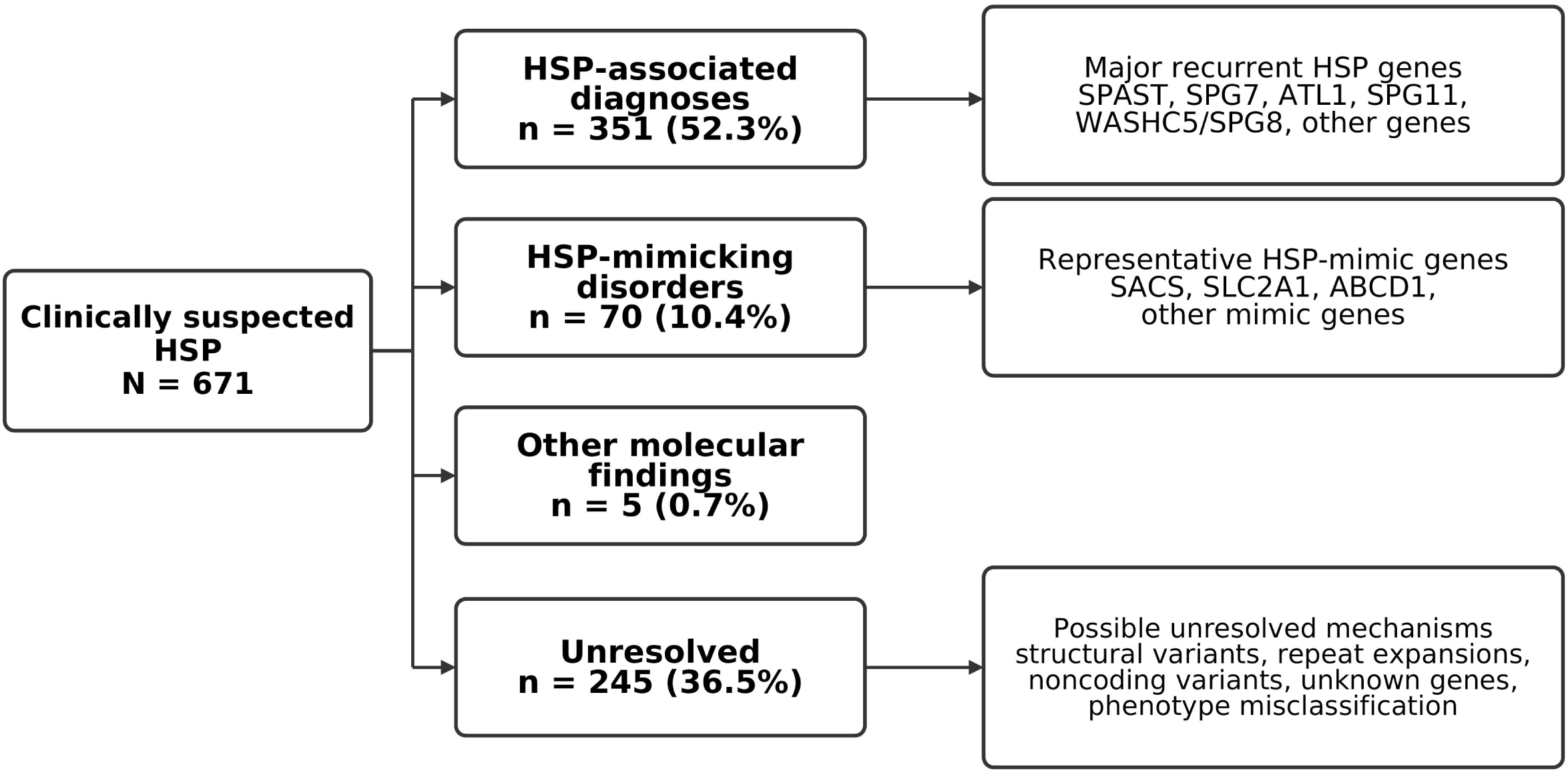
Diagnostic architecture of the CanHSP cohort (N=671). Mutually exclusive outcomes include HSP-associated, HSP-mimicking, other molecular, and unresolved findings. Percentages use N=671; listed genes and unresolved mechanisms are representative, not exhaustive.

Twenty-three of the 426 patients with molecular findings had findings in more than one distinct gene. Each of these individuals contributed once to the molecular-outcome count of 426, but contributed once to the frequency of each retained gene in gene-level analyses; consequently, gene-level counts were non-mutually exclusive. Among the 23, individuals with at least one HSP-associated gene were retained in the HSP-associated molecular finding group when the HSP-associated gene was considered the most plausible explanation for the phenotype. In the *AP5B1*–*PEX16* case, *AP5B1* was not retained as explanatory, and the individual was represented under *PEX16* alone in Supplementary Table S7, leaving 22 individuals in combined-gene rows across Supplementary Tables S5 and S7. In the full cohort’s inheritance patterns, 269/671 (40.1%) had autosomal dominant HSP (AD-HSP), 139/671 (20.7%) had autosomal recessive HSP (AR-HSP), and 21/671 (3.1%) had X-linked HSP (XL-HSP). The remaining 242/671 (36.1%) had no single clear inheritance category assigned and were labeled unknown.

We then examined patient-level gene frequencies among molecular findings. Table 2A and Table 2B includes 80 distinct genes classified as HSP-associated or HSP-mimic genes, including 36 HSP-associated genes. The most frequently identified genes with disease-causing variants were *SPAST* (151/671, 22.5%), *SPG7* (38/671, 5.7%), *ATL1* (35/671, 5.2%), *SPG11* (24/671, 3.6%), *SPG8* (22/671, 3.3%), *PLP1* (13/671, 1.9%), *ALS2* (8/671, 1.2%), *REEP1* (8/671, 1.2%), *CYP7B1* (7/671, 1.0%), and *KIF1A* (6/671, 0.9%) (Figure 2). Each remaining gene accounted for less than 1% individually, as shown in Table 2A and Table 2B.

**Table 2A.** Patient- and family-level frequencies of genes implicated in molecular findings in HSP.

| Gene | Associated condition | Patients, n | % of patients (N=671) | Families, n | % of families (N=423) |
| --- | --- | --- | --- | --- | --- |
| SPAST | SPG4 | 151 | 22.5% | 64 | 15.1% |
| SPG7 | SPG7 | 38 | 5.7% | 32 | 7.6% |
| ATL1 | SPG3A | 35 | 5.2% | 11 | 2.6% |
| SPG11 | SPG11 | 24 | 3.6% | 20 | 4.7% |
| WASHC5/SPG8 | SPG8 | 22 | 3.3% | 5 | 1.2% |
| PLP1 | SPG2 | 13 | 1.9% | 5 | 1.2% |
| ALS2 | Infantile-onset ascending hereditary spastic paralysis (IAHSP) | 8 | 1.2% | 2 | 0.5% |
| REEP1 | SPG31 | 8 | 1.2% | 8 | 1.9% |
| CYP7B1 | SPG5A | 7 | 1.0% | 6 | 1.4% |
| KIF1A | SPG30 | 6 | 0.9% | 5 | 1.2% |
| CAPN1 | SPG76 | 4 | 0.6% | 3 | 0.7% |
| ALDH18A1 | SPG9A/SPG9B | 3 | 0.4% | 1 | 0.2% |
| ATP13A2 | SPG78 | 3 | 0.4% | 3 | 0.7% |
| FA2H | SPG35 | 3 | 0.4% | 3 | 0.7% |
| BSCL2 | SPG17 | 2 | 0.3% | 1 | 0.2% |
| ERLIN2 | SPG18 | 2 | 0.3% | 1 | 0.2% |
| FARS2 | SPG77 | 2 | 0.3% | 1 | 0.2% |
| GBA2 | SPG46 | 2 | 0.3% | 1 | 0.2% |
| KIF5A | SPG10 | 2 | 0.3% | 2 | 0.5% |
| MARS | SPG70 | 2 | 0.3% | 2 | 0.5% |
| NIPA1 | SPG6 | 2 | 0.3% | 2 | 0.5% |
| PNPLA6 | SPG39 | 2 | 0.3% | 2 | 0.5% |
| SPTAN1 | SPTAN1-related neurodevelopmental/epileptic encephalopathy; HSP/ataxia phenotype reported | 2 | 0.3% | 2 | 0.5% |
| ZFYVE26 | SPG15 | 2 | 0.3% | 2 | 0.5% |
| AP4S1 | SPG52 | 1 | 0.1% | 1 | 0.2% |
| C19ORF12 | SPG43 | 1 | 0.1% | 1 | 0.2% |
| FICD | SPG92/ Infantile-onset diabetes mellitus coupled with severe neurodevelopmental delay | 1 | 0.1% | 1 | 0.2% |
| HSPD1 | SPG13 | 1 | 0.1% | 1 | 0.2% |
| KCNA2 | KCNA2-related epileptic encephalopathy/ataxia; rare HSP phenotype reported | 1 | 0.1% | 1 | 0.2% |
| MAG | SPG75 | 1 | 0.1% | 1 | 0.2% |
| NT5C2 | SPG45 | 1 | 0.1% | 1 | 0.2% |
| REEP2 | SPG72 | 1 | 0.1% | 1 | 0.2% |

| Gene | Associated condition | Patients, n | % of patients<br>(N=671) | Families, n | % of families<br>(N=423) |
| --- | --- | --- | --- | --- | --- |
| SLC16A2 | SPG22 | 1 | 0.1% | 1 | 0.2% |
| TECPR2 | SPG49 | 1 | 0.1% | 1 | 0.2% |
| UBAP1 | SPG80 | 1 | 0.1% | 1 | 0.2% |
| ZFYVE27 | SPG33 | 1 | 0.1% | 1 | 0.2% |

Counts represent the number of patients or families with a molecular finding involving each gene and are non-mutually exclusive. An individual with findings involving more than one gene contributes once to each implicated gene. Accordingly, gene-level counts must not be summed to obtain the number of molecularly diagnosed individuals. Tables 2A and 2B include HSP-associated and HSP-mimic genes only. The 421 patients in the HSP-associated and HSP-mimic groups contribute 443 gene assignments across Tables 2A and 2B.

**Table 2B.**
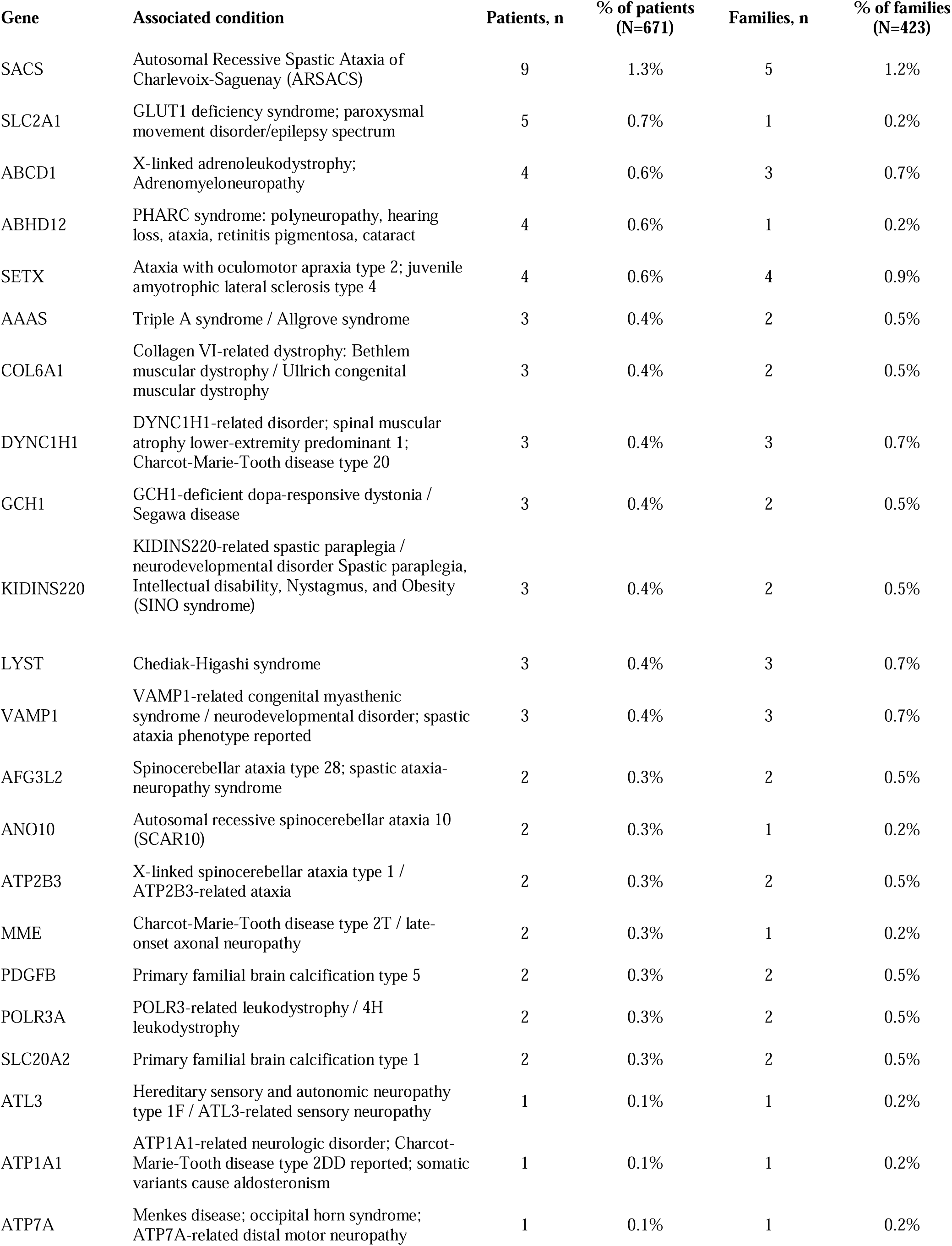
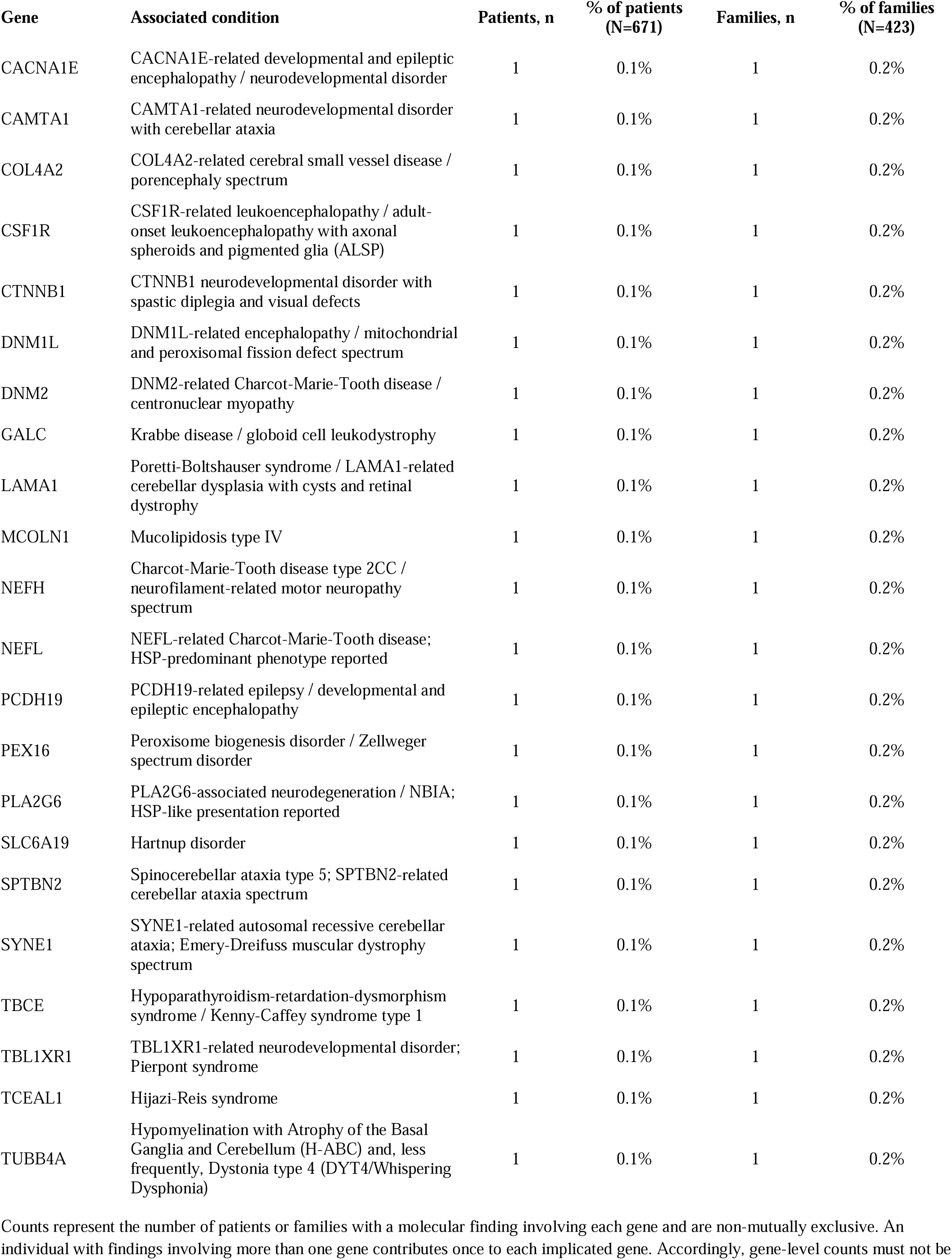
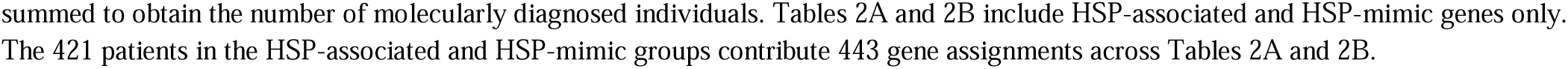
Patient- and family-level frequencies of genes implicated in molecular findings in HSP mimics.

**Figure 2.**
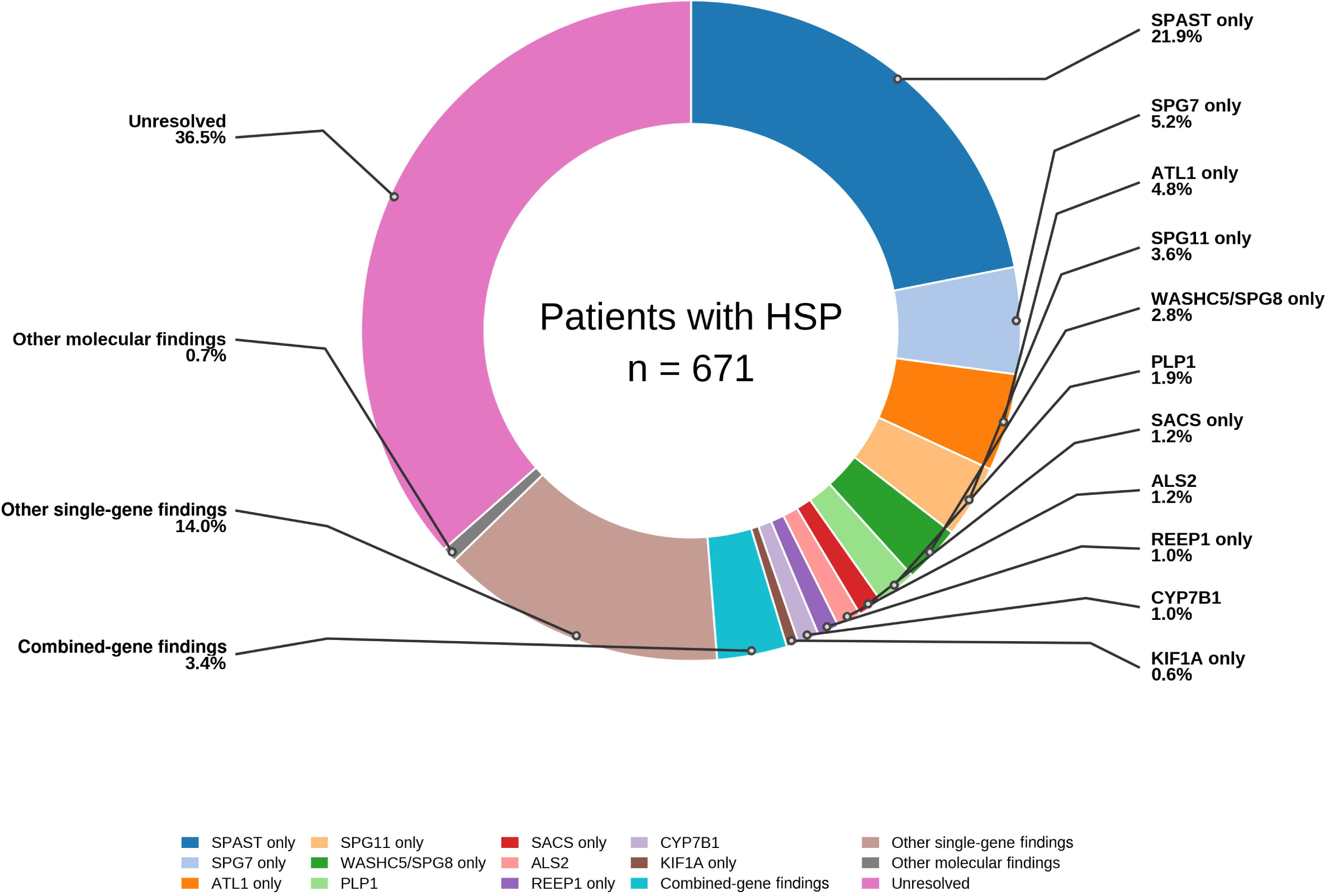
Molecular findings groups in CanHSP (N=671). Recurrent-gene slices are single-gene findings; combined-gene findings are separate. Remaining HSP-associated and HSP-mimic single-gene findings are grouped as “Other single-gene findings.” Gene-level frequencies are reported in Table 2A and Table 2B.

Family-level analysis showed that some recurrent genes were concentrated within a small number of pedigrees. WASHC5/SPG8, for example, was identified in 22 individuals from five families, including 10 individuals from a single family, whereas SPAST was identified in 151 individuals distributed across 64 families. Thus, patient-level frequencies for some genes partly reflect within-family clustering.

#### Clinical characterization of the major recurrent genes

We next examined the clinical profiles of the five major recurrent genes (Table 3). Because of the overlap from combined-gene findings, genotype-phenotype summaries were restricted to the corresponding single-gene molecular finding groups in Supplementary Table S5. For each clinical feature, percentages were calculated only among patients in the relevant single-gene finding group with available data for that feature.

**Table 3.**
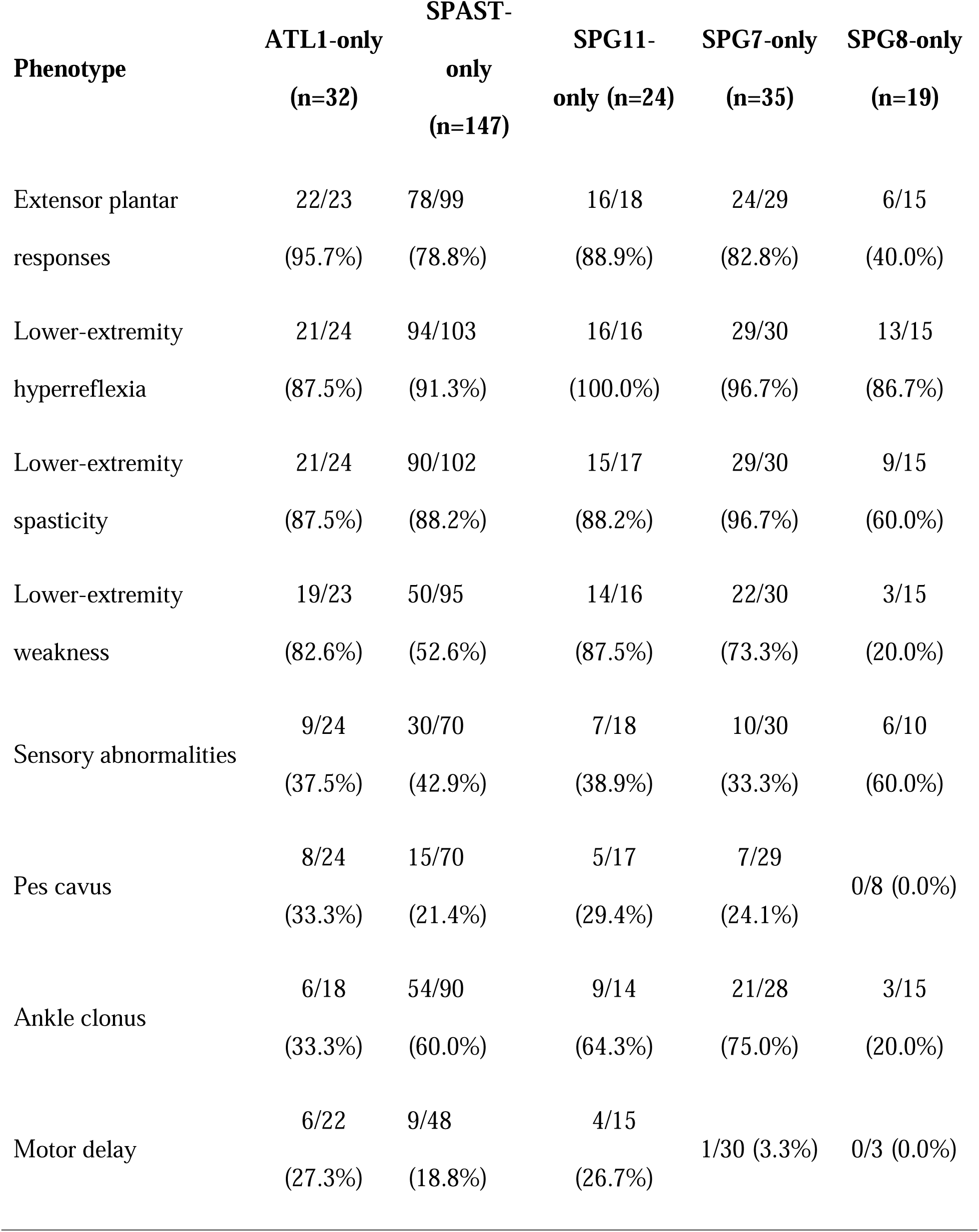
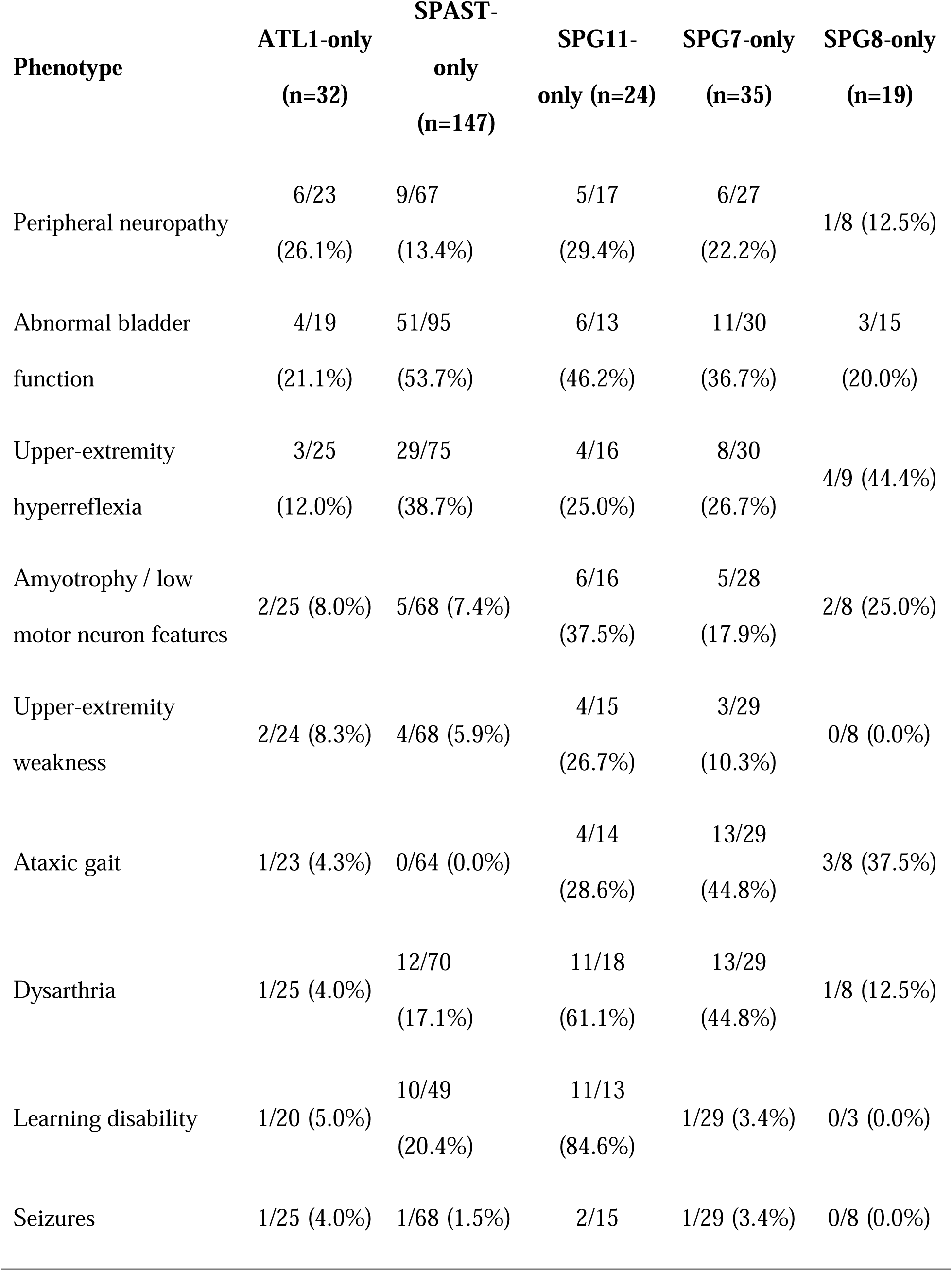
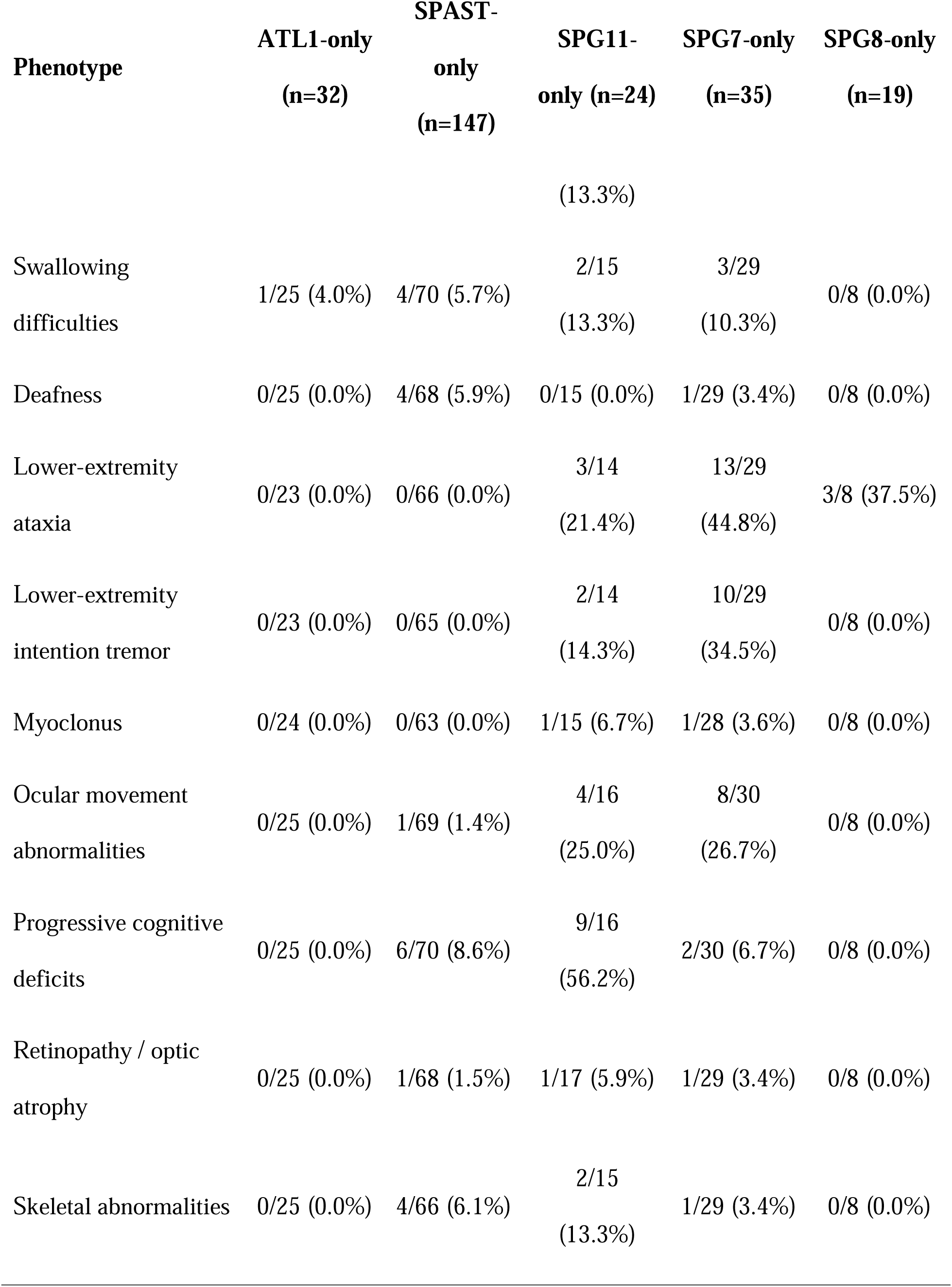
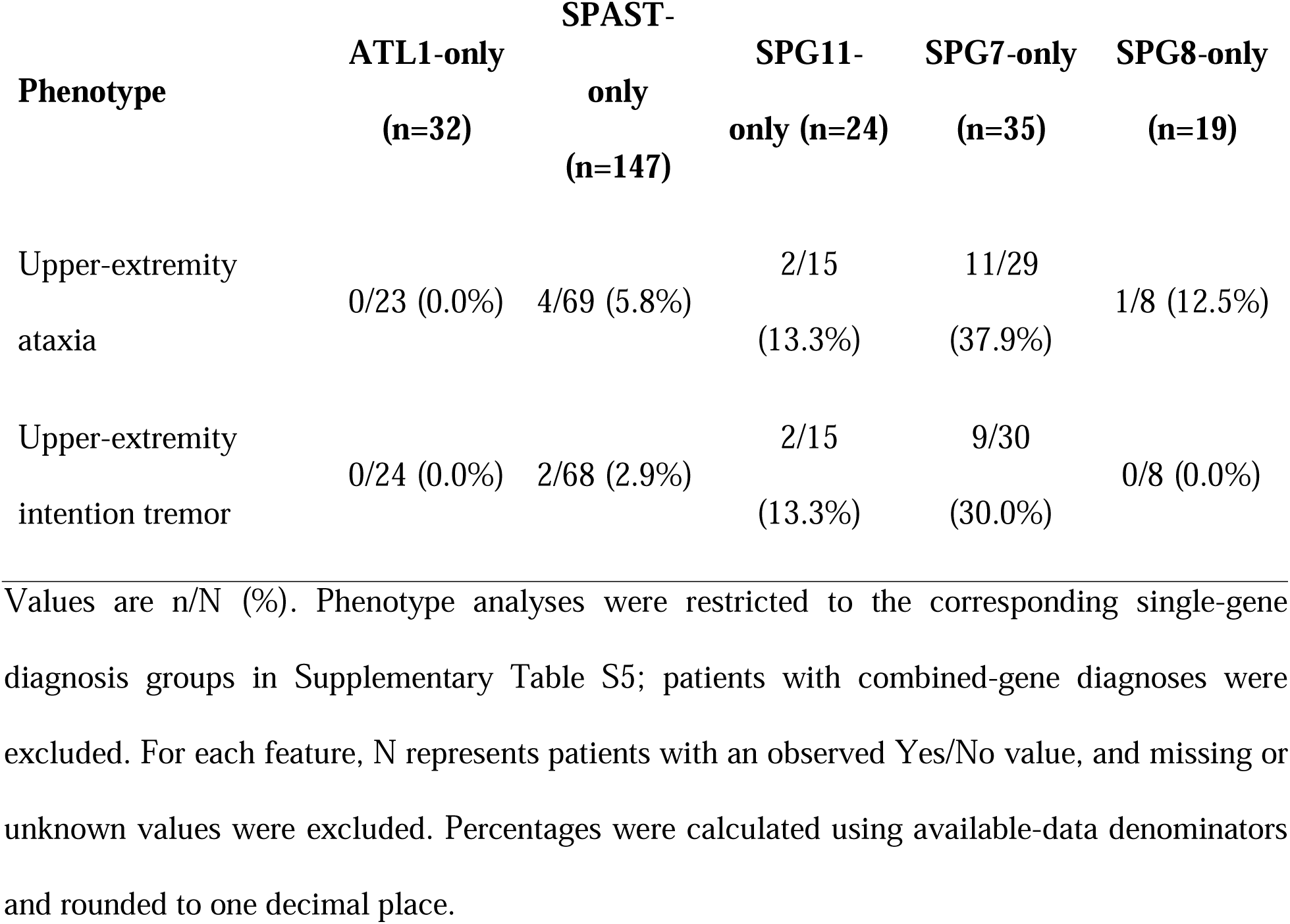
Clinical phenotype summary of major single-gene molecular diagnosis groups.

| **Phenotype** | **ATL1-only**<br>**(n=32)** | **SPAST-**<br>**only**<br>**(n=147)** | **SPG11-**<br>**only (n=24)** | **SPG7-only**<br>**(n=35)** | **SPG8-only**<br>**(n=19)** |
| --- | --- | --- | --- | --- | --- |
| Extensor plantar responses | 22/23<br>(95.7%) | 78/99<br>(78.8%) | 16/18<br>(88.9%) | 24/29<br>(82.8%) | 6/15<br>(40.0%) |
| Lower-extremity hyperreflexia | 21/24<br>(87.5%) | 94/103<br>(91.3%) | 16/16<br>(100.0%) | 29/30<br>(96.7%) | 13/15<br>(86.7%) |
| Lower-extremity spasticity | 21/24<br>(87.5%) | 90/102<br>(88.2%) | 15/17<br>(88.2%) | 29/30<br>(96.7%) | 9/15<br>(60.0%) |
| Lower-extremity weakness | 19/23<br>(82.6%) | 50/95<br>(52.6%) | 14/16<br>(87.5%) | 22/30<br>(73.3%) | 3/15<br>(20.0%) |
| Sensory abnormalities | 9/24<br>(37.5%) | 30/70<br>(42.9%) | 7/18<br>(38.9%) | 10/30<br>(33.3%) | 6/10<br>(60.0%) |
| Pes cavus | 8/24<br>(33.3%) | 15/70<br>(21.4%) | 5/17<br>(29.4%) | 7/29<br>(24.1%) | 0/8 (0.0%) |
| Ankle clonus | 6/18<br>(33.3%) | 54/90<br>(60.0%) | 9/14<br>(64.3%) | 21/28<br>(75.0%) | 3/15<br>(20.0%) |
| Motor delay | 6/22<br>(27.3%) | 9/48<br>(18.8%) | 4/15<br>(26.7%) | 1/30 (3.3%) | 0/3 (0.0%) |
| Peripheral neuropathy | 6/23<br>(26.1%) | 9/67<br>(13.4%) | 5/17<br>(29.4%) | 6/27<br>(22.2%) | 1/8 (12.5%) |
| Abnormal bladder<br>function | 4/19<br>(21.1%) | 51/95<br>(53.7%) | 6/13<br>(46.2%) | 11/30<br>(36.7%) | 3/15<br>(20.0%) |
| Upper-extremity<br>hyperreflexia | 3/25<br>(12.0%) | 29/75<br>(38.7%) | 4/16<br>(25.0%) | 8/30<br>(26.7%) | 4/9 (44.4%) |
| Amyotrophy / low<br>motor neuron features | 2/25 (8.0%) | 5/68 (7.4%) | 6/16<br>(37.5%) | 5/28<br>(17.9%) | 2/8 (25.0%) |
| Upper-extremity<br>weakness | 2/24 (8.3%) | 4/68 (5.9%) | 4/15<br>(26.7%) | 3/29<br>(10.3%) | 0/8 (0.0%) |
| Ataxic gait | 1/23 (4.3%) | 0/64 (0.0%) | 4/14<br>(28.6%) | 13/29<br>(44.8%) | 3/8 (37.5%) |
| Dysarthria | 1/25 (4.0%) | 12/70<br>(17.1%) | 11/18<br>(61.1%) | 13/29<br>(44.8%) | 1/8 (12.5%) |
| Learning disability | 1/20 (5.0%) | 10/49<br>(20.4%) | 11/13<br>(84.6%) | 1/29 (3.4%) | 0/3 (0.0%) |
| Seizures | 1/25 (4.0%) | 1/68 (1.5%) | 2/15 | 1/29 (3.4%) | 0/8 (0.0%) |
|  |  |  | (13.3%) |  |  |
| Swallowing difficulties | 1/25 (4.0%) | 4/70 (5.7%) | 2/15<br>(13.3%) | 3/29<br>(10.3%) | 0/8 (0.0%) |
| Deafness | 0/25 (0.0%) | 4/68 (5.9%) | 0/15 (0.0%) | 1/29 (3.4%) | 0/8 (0.0%) |
| Lower-extremity ataxia | 0/23 (0.0%) | 0/66 (0.0%) | 3/14<br>(21.4%) | 13/29<br>(44.8%) | 3/8 (37.5%) |
| Lower-extremity intention tremor | 0/23 (0.0%) | 0/65 (0.0%) | 2/14<br>(14.3%) | 10/29<br>(34.5%) | 0/8 (0.0%) |
| Myoclonus | 0/24 (0.0%) | 0/63 (0.0%) | 1/15 (6.7%) | 1/28 (3.6%) | 0/8 (0.0%) |
| Ocular movement abnormalities | 0/25 (0.0%) | 1/69 (1.4%) | 4/16<br>(25.0%) | 8/30<br>(26.7%) | 0/8 (0.0%) |
| Progressive cognitive deficits | 0/25 (0.0%) | 6/70 (8.6%) | 9/16<br>(56.2%) | 2/30 (6.7%) | 0/8 (0.0%) |
| Retinopathy / optic atrophy | 0/25 (0.0%) | 1/68 (1.5%) | 1/17 (5.9%) | 1/29 (3.4%) | 0/8 (0.0%) |
| Skeletal abnormalities | 0/25 (0.0%) | 4/66 (6.1%) | 2/15<br>(13.3%) | 1/29 (3.4%) | 0/8 (0.0%) |
| Upper-extremity<br>ataxia | 0/23 (0.0%) | 4/69 (5.8%) | 2/15<br>(13.3%) | 11/29<br>(37.9%) | 1/8 (12.5%) |
| Upper-extremity<br>intention tremor | 0/24 (0.0%) | 2/68 (2.9%) | 2/15<br>(13.3%) | 9/30<br>(30.0%) | 0/8 (0.0%) |

Values are n/N (%). Phenotype analyses were restricted to the corresponding single-gene diagnosis groups in Supplementary Table S5; patients with combined-gene diagnoses were excluded. For each feature, N represents patients with an observed Yes/No value, and missing or unknown values were excluded. Percentages were calculated using available-data denominators and rounded to one decimal place.

*SPAST* variants (SPG4) were the most common, occurring in 151 patients (35.4% of patients with molecular findings; 22.5% of the total cohort). Among 147 patients with a *SPAST*-only molecular finding, 116/147 (78.9%) were classified as pure HSP. Dominant pyramidal features included lower-extremity hyperreflexia (94/103, 91.3%), spasticity (90/102, 88.2%), extensor plantar responses (78/99, 78.8%), and lower-extremity weakness (50/95, 52.6%). Cerebellar manifestations were uncommon: ataxic gait was absent in 64 patients with available data and lower-extremity ataxia in 66, while upper-extremity ataxia occurred in 4/69 (5.8%) and ocular movement abnormalities in 1/69 (1.4%).

Biallelic *SPG7* variants were the second most common cause of HSP, identified in 38 patients (8.9% of patients with molecular findings; 5.7% of the total cohort). Among 35 patients with an *SPG7*-only finding, 19/35 (54.3%) had a complicated phenotype, broader than that seen with *SPAST*. Pyramidal signs included lower-extremity hyperreflexia (29/30, 96.7%), spasticity (29/30, 96.7%), extensor plantar responses (24/29, 82.8%), and weakness (22/30, 73.3%). Cerebellar and bulbar features included ataxic gait and lower-extremity ataxia (13/29, 44.8% each), upper-extremity ataxia (11/29, 37.9%), lower-extremity intention tremor (10/29, 34.5%), upper-extremity intention tremor (9/30, 30.0%), and dysarthria (13/29, 44.8%). Abnormal bladder function occurred in 11/30 (36.7%) and ankle clonus in 21/28 (75.0%).

*ATL1* variants were the second most common AD-HSP (SPG3A) cause, identified in 35 patients (8.2% of patients with molecular findings; 5.2% of the total cohort). Among 32 *ATL1*-only findings, 24/32 (75.0%) were pure HSP. *ATL1* was characterized by early pyramidal involvement: lower-extremity hyperreflexia and spasticity each occurred in 21/24 (87.5%), lower-extremity weakness in 19/23 (82.6%), and extensor plantar responses in 22/23 (95.7%). Cerebellar and bulbar manifestations were uncommon compared with *SPG7* and *SPG11*: dysarthria occurred in 1/25 (4.0%), ataxic gait in 1/23 (4.3%), upper-extremity weakness in 2/24 (8.3%), and amyotrophy or lower motor neuron features in 2/25 (8.0%).

Biallelic *SPG11* variants were identified in 24 patients (5.6% of patients with molecular findings; 3.6% of the total cohort). All 24 were single-gene findings, with 15/24 (62.5%) classified as complicated HSP. *SPG11* showed the most complicated clinical profile among the major recurrent genes. Pyramidal features included lower-extremity hyperreflexia (16/16, 100.0%), spasticity (15/17, 88.2%), weakness (14/16, 87.5%), and extensor plantar responses (16/18, 88.9%). Developmental, cognitive, and lower motor neuron features included learning disability (11/13, 84.6%), progressive cognitive deficits (9/16, 56.2%), dysarthria (11/18, 61.1%), and amyotrophy or lower motor neuron features (6/16, 37.5%). Additional manifestations included ataxic gait (4/14, 28.6%), ocular movement abnormalities (4/16, 25.0%), and peripheral neuropathy (5/17, 29.4%).

*SPG8* variants were the fifth most frequent cause of HSP, identified in 22 patients (5.2% of patients with molecular findings; 3.3% of the total cohort). Among 19 *SPG8*-only findings, 15/19 (78.9%) were pure HSP. Compared with *SPAST* and *ATL1*, *SPG8* showed a largely pyramidal but more variable phenotype. Lower-extremity hyperreflexia occurred in 13/15 (86.7%), spasticity in 9/15 (60.0%), extensor plantar responses in 6/15 (40.0%), and lower-extremity weakness in 3/15 (20.0%). Sensory abnormalities occurred in 6/10 (60.0%). Among patients with available cerebellar assessments, ataxic gait and lower-extremity ataxia each occurred in 3/8 (37.5%), and upper-extremity ataxia in 1/8 (12.5%).

#### HSP mimics

Of the 426 individuals with molecular findings, 70 (16.4%; 10.4% of the full cohort) had HSP-mimicking molecular findings in genes usually not associated with HSP despite a clinically HSP-like presentation (Supplementary Table S7). These included hereditary ataxias, metabolic disorders, neuropathy or neuromuscular disorders, and multisystem syndromic conditions (Table 2B). The most common HSP-mimic gene in our population was *SACS*, known for causing Autosomal Recessive Spastic Ataxia of Charlevoix-Saguenay (ARSACS), reflecting the overlap between HSP and ARSACS in the French-Canadian population in Quebec ^35^. Among patients with HSP mimics and available clinical data, lower-extremity spasticity, lower-extremity hyperreflexia, and extensor plantar responses were present in 54/59, 51/56, and 47/56 patients, respectively, while lower-extremity weakness was present in 41/56. Additional features reflected overlap with ataxic, neuropathic, neurodevelopmental, and leukodystrophy-like disorders, including motor delay (17/43), dysarthria (15/48), ataxic gait (14/42), sensory abnormalities (14/43), learning disability (15/44), and lower-extremity ataxia (12/44). Abnormal brain and spine MRI findings were reported in 13/54 and 13/54 patients with available imaging data, respectively, and phenotype classification remained relatively balanced between pure and complicated presentations (40/70 and 30/70).

#### Candidate Novel Variants

We identified four rare variants for further evaluation (Supplementary Table S8). Three variants, in PEX16, AFG3L2, and TUBB4A, were absent from the queried population and disease databases and were not found in previous literature; these were retained as candidate novel variants. The *KIDINS220* c.4148A>G variant was absent from all assessed databases except ClinVar where it is reported as a variant of uncertain significance; therefore, it was considered rare but not completely novel.

## Discussion

This multicenter Canadian cohort demonstrates substantial genetic heterogeneity among individuals with clinically suspected HSP. The 63.5% molecular finding rate reflects findings accumulated through multiple testing pathways, principally gene-panel testing and WES, with CNV/MLPA assessment when available, rather than a uniform yield from a single strategy. These findings support flexible genetic testing and continued reanalysis ^1,3,7,18,27^.

*SPAST* was the most frequent gene, followed by *SPG7*, *ATL1*, *SPG11*, and *SPG8*, but many additional genes were observed in small numbers of patients. The comparatively high patient-level frequency of WASHC5/SPG8 cases should therefore be interpreted in the context of pedigree structure. Although clustering within a small number of families may be compatible with population-specific enrichment, we did not perform haplotype analysis and therefore cannot establish a founder effect. A recurrent-gene panel would capture many findings, especially SPG4, but would miss many rare HSP-associated and HSP mimic findings, supporting broader sequencing approaches ^10–12,36^. This long-tail distribution also highlights the continuing challenge of variant interpretation in HSP, particularly when rare variants are identified in genes with variable inheritance patterns, overlapping phenotypes, or evolving evidence for disease association ^15,21,32^.

The observation of molecular findings involving more than one clinically plausible gene raises the possibility of non-Mendelian contributions to the phenotype, particularly in HSP, where genetic heterogeneity, variable expressivity, and overlapping neurogenetic presentations are common. This possibility has been highlighted in SPG7-associated disease and may also be relevant to other HSP-related genes ^20^. However, without systematic segregation, zygosity, phasing, and functional data, these observations should be interpreted cautiously and considered hypothesis-generating rather than definitive evidence of oligogenic inheritance.

The unresolved fraction highlights limitations of panel testing and short-read exome sequencing in HSP ^10,12–14^. These approaches effectively detect coding single-nucleotide variants and small insertions or deletions but are less suited to structural variants, repeat expansions, complex or difficult-to-map regions, and non-coding or deep intronic variants. Long-read sequencing can complement conventional testing by spanning repetitive or structurally complex regions, improving structural-variant detection and long-range phasing, and resolving complex alleles. Studies of unresolved rare-disease cohorts support its use as a second-line approach after negative conventional testing ^37,38^.

HSP mimic-associated findings are clinically important because these disorders may resemble HSP yet differ in prognosis, counseling, surveillance, and management. Mimic findings therefore required genotype-phenotype concordance rather than simply a variant in a non-HSP gene. Overlap with hereditary ataxias, leukodystrophies, neuropathies, and metabolic or mitochondrial diseases supports comprehensive testing and phenotypic reassessment when presentations are atypical or additional neurological features emerge.

Complicated HSP was less often familial, had a higher relative contribution of autosomal recessive and X-linked inheritance, and was associated with greater disease severity and disability than pure HSP, although disease duration, referral patterns, and missing clinical data may have influenced these comparisons^27,39^.

This study has several limitations. Recruitment spanned decades during which phenotyping, sequencing technologies, and annotation evolved, with non-uniform annotation quality across resolved cases. Thus, the molecular finding rate reflects findings documented in registry records through multiple pathways rather than a standardized pipeline. Original case-level documentation, including exon-level MLPA reports, was unavailable for all CNV records; these cases were classified as unresolved, excluded from solved gene-frequency estimates, and not reported as a separate structural-variant subgroup. HSP-associated versus HSP mimic classification relied on recorded gene-disease associations and phenotype compatibility, with case-level validation limited by registry data. Phenotypic data were incomplete and potentially non-randomly missing, particularly in older entries. Related individuals reduce patient-level independence, although family-level counts were provided; referral-center ascertainment may also bias referral patterns. Finally, panel testing and WES rather than WGS may miss non-coding, deep intronic, copy-number or other structural variants, and repeat expansions. Nevertheless, the cohort’s size, national reach, and molecular characterization provide a strong overview of HSP genetics in Canada.

Recurrent genes such as *SPAST*, *SPG7* and *SPG11* account for a substantial proportion of findings, but extensive genetic heterogeneity remains a defining feature of the disorder. Continued expansion of genomic testing, variant curation, and gene discovery will be essential to improve diagnostic precision and clinical care in HSP.

## 4. Author contributions

SN conducted the analysis, interpreted the results, and drafted the manuscript. MAE conducted the original genetic analyses underlying the cohort dataset, contributed to interpretation of the results, and critically revised the manuscript. HY assisted with the analysis. EY, SA, and JA provided expertise and support. SDS, KMB, MT, OS, GY, and ND recruited the participants and collected data. ZG-O and GAR supervised the study and contributed to interpretation of the results. All authors reviewed, edited, and approved the final manuscript. SN and MAE contributed equally to this work.

## 5. Statements and declarations

### 5.1. Ethical considerations

Ethics approval for the research study was granted by the McGill University Research Ethics Board.

### 5.2. Consent to participate

Informed consent forms were signed by all participants before entering the study.

### 5.3. Consent for publication

Not applicable.

### 5.6. Data availability

All data produced in the present study are available at reasonable request to the authors.

## 6. Acknowledgements

We would like to thank the patients and their families for participating in this study. We thank the members of CanHSP who made this study possible, including Meredith Gillespie. G.A.R. holds the Wilder Penfield Chair in Neurosciences. Z.G.O. is supported by the Fonds de recherche du Québec–Santé Chercheur-Boursier award and is a William Dawson Scholar.

## Funding agencies

This study received funding from the CIHR Emerging Team Grant in partnership with the Canadian Organization for Rare Disorders (CORD) (grant no. RN127580– 260005), as well as from a CIHR Foundation Grant awarded to G.A.R.

