## Supplemental Figures for "THE GENETIC LANDSCAPE OF HEREDITARY SPASTIC PARAPLEGIA IN CANADA": Supplementary figures.docx

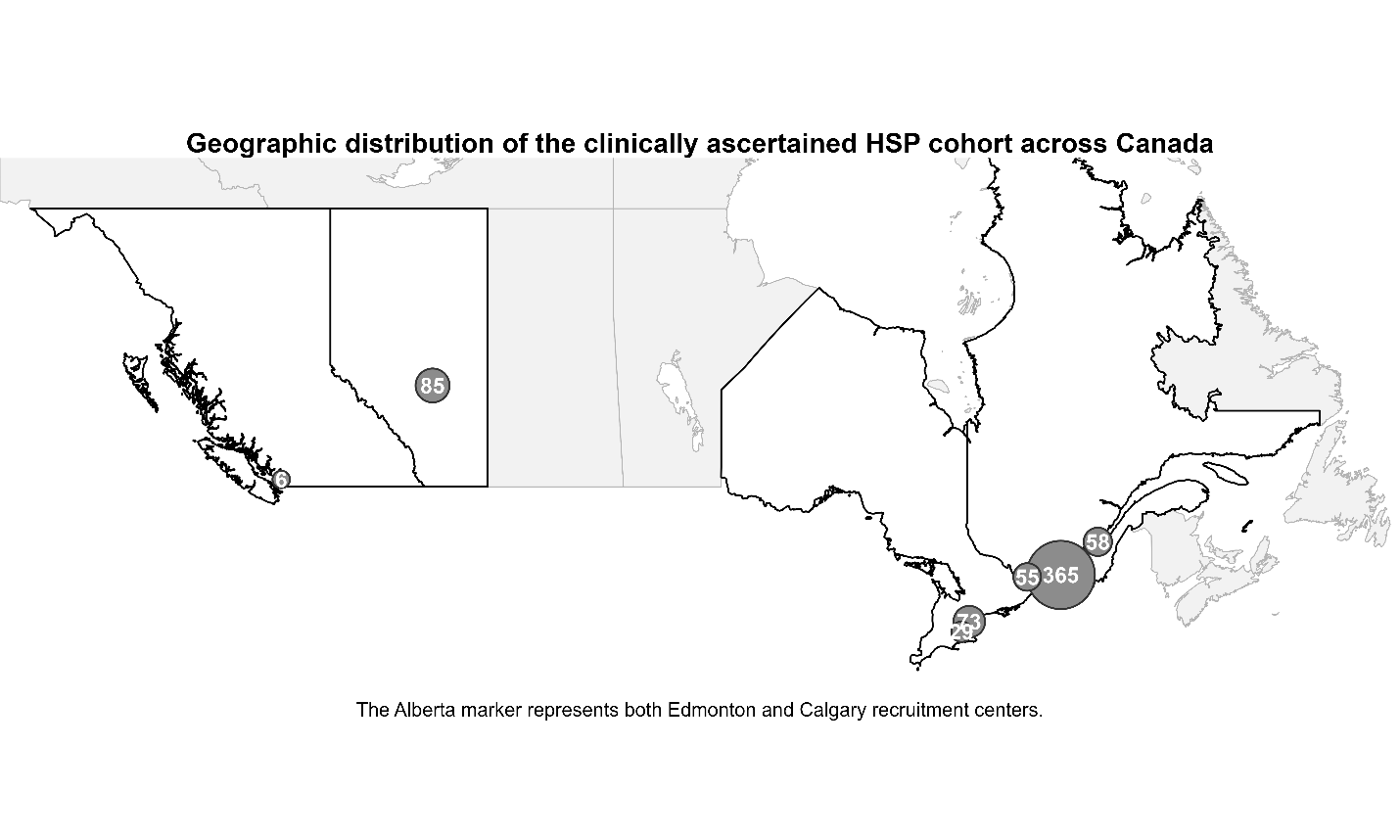

**Supplementary** **Figure S1**. **Geographic distribution of the clinically ascertained HSP cohort across Canada (N = 671)**. Circles indicate the number of affected patients suspected for HSP recruited at participating Canadian clinical centers. HSP, hereditary spastic paraplegia.

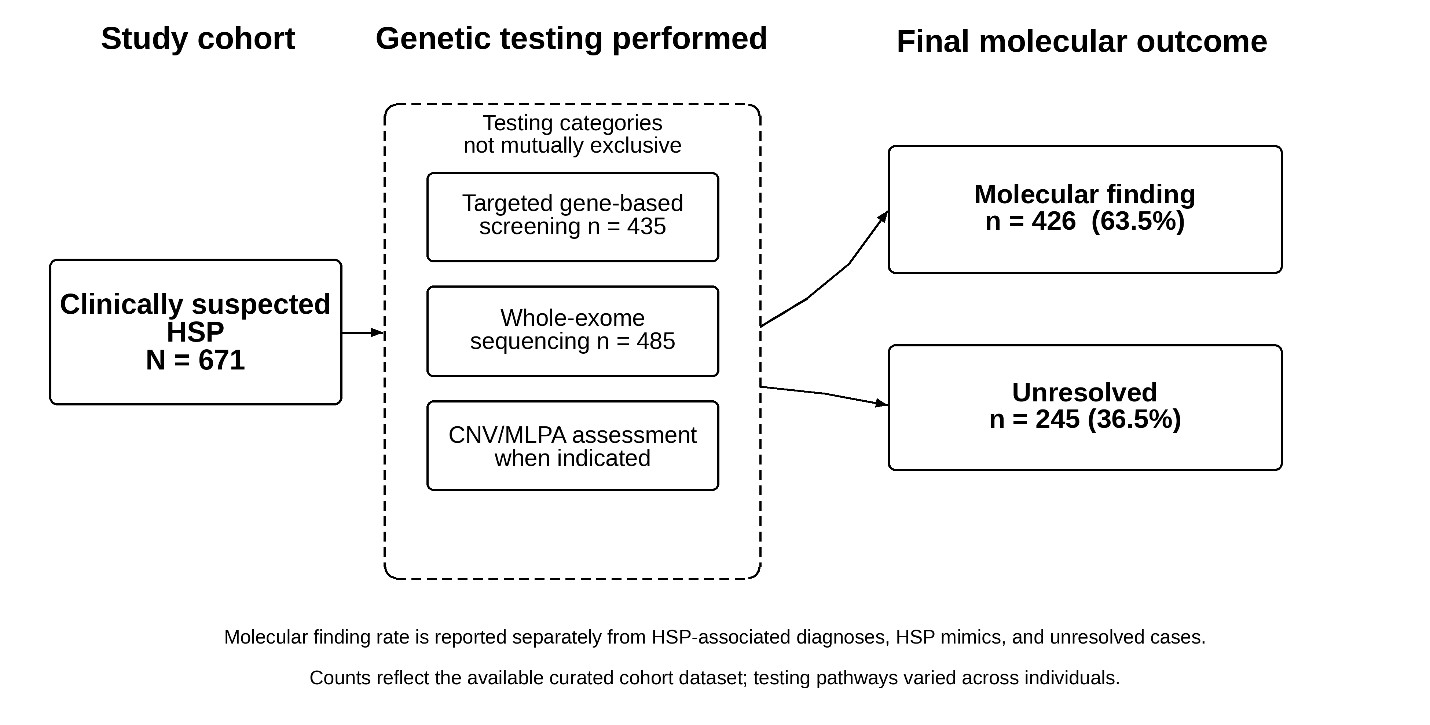

**Supplementary** **Figure S2. CanHSP cohort testing overview and molecular outcome.** Overview of genetic testing and final molecular outcomes in 671 clinically suspected HSP cases. Testing categories are non-mutually exclusive; final molecular outcomes are mutually exclusive and sum to N=671.
